# Bone Recovery After Treatment Study (BRATS) - A Protocol for a Prospective, Observational Study in Multiple Myeloma

**DOI:** 10.64898/2026.01.20.26344424

**Authors:** R.E. Andrews, I. Jolly, J.E. Brown, M.A. Lawson, A.D. Chantry

**Affiliations:** School of Medicine and Population Health, and Mellanby Centre for Musculoskeletal Research, Medical School, Beech Hill Road, Sheffield, S10 2RX, UK; Sheffield Teaching Hospitals NHS Foundation Trust, Royal Hallamshire Hospital, Sheffield S10 2JF, UK

**Keywords:** multiple myeloma, bone disease, osteolytic lesions, bone anabolic, quality of life, bone marrow recovery

## Abstract

Cancer-induced bone disease is a huge burden on patient lives and costs the NHS millions of pounds every year. Breast, prostate and lung cancer can all lead to poor skeletal outcomes, but patients particularly at risk are those with a diagnosis of multiple myeloma (1). Despite response to tumour targeting treatments, patients experience debilitating bone pain and fractures, affecting quality of life (2, 3). Currently, myeloma patients who are eligible, are offered treatment with induction chemotherapy followed by autologous stem cell transplant (ASCT). Most eligible patients also receive bisphosphonates, to reduce skeletal morbidity, but this treatment is not optimal, or even conducive for bone recovery. Therefore, we wanted to assess whether current induction chemotherapy regimens have the capacity to reset the bone marrow microenvironment (BMME).

This prospective observational cohort study will recruit newly diagnosed myeloma patients from Sheffield Teaching Hospitals NHS Foundation Trust. Ethical approval has been granted to undergo two recruitment periods; cohort 1 (20 participants, forming a pilot study) and cohort 2 (up to 100 participants with a streamlined follow-up design).

Macro-architectural skeletal bone disease will be assessed by whole-body low-dose CT (WBLDCT) scans, in which osteolytic lesions will be monitored longitudinally. Micro-architecture will be assessed by micro-CT scanning bone marrow trephine samples, and analysing changes in trabecular bone. Bone integrity will be assessed using computational models of both whole body skeletal and micro trephine images. Fasting serum samples will be collected to assess changes in bone turnover markers. This will be supported by histomorphometry and immunohistochemistry analysis of trephine sections. All samples / imaging will be performed at baseline and follow-up. Monitoring of quality of life (validated questionnaires) and occurrence of skeletal related events (SREs) will also take place. The observational period will end one year post ASCT. Data collected from this study, will provide an invaluable opportunity to comprehensively assess myeloma-induced bone disease and broaden our understanding of the disease course. It may also prove a valuable resource to guide the design of interventional clinical studies exploring novel bone-targeted therapies, including bone anabolic therapeutics, moving forward.

## Background

Myeloma bone disease (MBD) is a substantial complication, affecting up to 90% of patients with myeloma (1). Despite tumour depleting therapies, patients are often left with considerable bone pain, poor mobility, and reduced functional and economic capacity. Many patients report reduced quality of life (QOL) because of sub-optimally treated bone disease (2, 3). The pathophysiology of osteolytic lesion development has been better characterised in recent years. The basic precedent being that the presence of tumour cells in the BMME causes chemokines/cytokines release, causing an overstimulation of osteoclast activity and inhibition of osteoblasts, leading to a net bone loss through unopposed osteoclastic bone resorption (4). Recent studies have also suggested that osteocytes, the main orchestrators of bone remodelling, are also dramatically affected by the presence of myeloma cells. This basis (although simplified for the purpose of this text), underpins the current developments of novel bone-targeted therapies in the field of cancer-induced bone pathology. Current preclinical data suggests that the catabolic state of bone is reset following treatment, hence giving a unique opportunity to introduce novel anabolic agents at this time point (5, 6). Therefore, we need to assess whether there is clinical evidence that the BMME can recover, and how best to support the recovery of existing bone disease for patients with myeloma.

Currently, little is understood about how the BMME behaves following current induction chemotherapy (plus bisphosphonates). Can the bone environment reset? Can osteoblast cells recover activity? Do all patients have the same potential to repair bone? Addressing these research questions will allow us to develop the best strategies for optimising future treatments in bone-targeted therapies.

To answer some of these questions, we assessed skeletal macro and micro architectural changes, bone turnover markers, histomorphology of bone marrow trephine samples and reported SREs / patient reported QOL questionnaires to gain in depth knowledge about how MBD behaves following induction chemotherapy. To the best of our knowledge, an in depth observational study of this nature has not previously been undertaken. This research will broaden our understanding of MBD and potentially aid the development of new bone-targeting therapies in the future. It will also help us to determine the best timeframe for the introduction of upcoming novel pharmacological treatments, such as bone anabolic agents. The study will also be relevant to other oncological conditions that have a high affinity to develop bone metastasis, such as breast and prostate cancer.

## Research Methods and Analyses

### Study Design and Setting

This is a prospective, observational study in a cohort of patients undergoing investigations for a suspected new diagnosis of myeloma. It is a single-centre study, recruiting patients at the Royal Hallamshire Hospital (Sheffield Teaching Hospitals NHS Foundation Trust, Sheffield, UK (STH)). There will be two recruitment cohorts: cohort 1 (20 participants, forming a pilot study) and cohort 2 (up to 100 participants with a streamlined follow-up design). Following the initial findings and analysis of cohort 1 (pilot study), adaptations may be made to the study design prior to cohort 2 (Fig. 1). Such adaptions may include reducing timepoints of outcome markers (if the pilot can identify the follow-up timepoints for optimal study readouts), as well as optimising choice of endpoints (including powering the second recruitment to a primary or composite endpoint that could be adapted to interventional trials in the future). All potential adaptations to the study design for recruitment of cohort 2 will be discussed with the study sponsor and submitted to the ethics board for consideration as a study amendment prior to opening phase 2 recruitment (Fig. 1).

**Figure 1.**
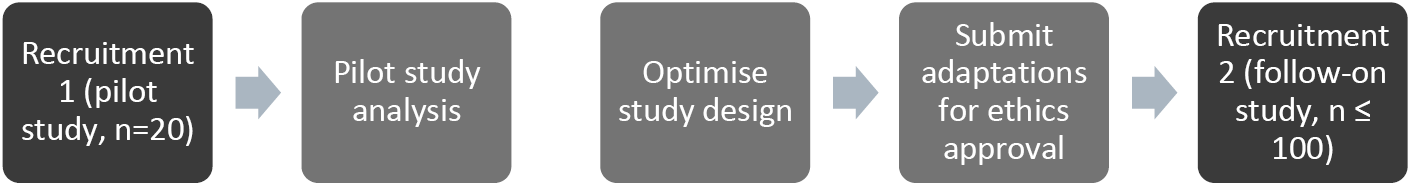
Study phases. The initial recruitment window (recruitment cohort 1) is a small pilot study (n=20), which will be followed by a period of analysis and study design optimisation, before the recruitment of cohort 2 (n≤100). The second study (cohort 2) will be streamlined optimising the best read-outs and endpoints to answer our research aims based on lessons learnt from the pilot study.

All research investigations, including obtaining research study samples, will take place within STH. Sample analysis will be performed at University of Sheffield (Sheffield, UK). Patients were consulted during conceptualization, to confirm the study’s relevance and acceptability to myeloma patients.

### Study Aim

Key research questions include:

1. Following induction chemotherapy, does the bone microenvironment recover adequately to facilitate the re-cultivation of healthy bone remodelling?
2. Can we capture evidence of bone recovery using comparative endpoints?
3. Can we identify an optimum time point for bone anabolic therapy in patients with MBD?

The primary aim of the study is to comprehensively assess changes in bone health in patients with MBD following first-line treatment (+/-ASCT). Based on observations in pre-clinical models of MBD, we anticipate skeletal changes following first line treatment (5, 6).

### Study Population

Patients undergoing a diagnostic bone marrow biopsy for suspected myeloma, also anticipated to be eligible for intensive first line treatment, will be invited to participate in the study. Patients will be given information about the study by their medical team during medical consultations. Inclusion and exclusion criteria for the study are listed in Table 1. Enrolled patients receiving a diagnosis of myeloma will continue with study follow-up for up to 12 months post first line treatment (either at engraftment post ASCT or following final cycle of intensive chemotherapy if ASCT did not take place) (Fig. 2). Patients will be advised that they can withdraw from the study at any point; however, previously collected data and tissue will be retained.

**Table 1.** Summary of eligibility criteria for study participation.

| Inclusion | Exclusion |
| --- | --- |
| • Age $\geq 18$<br>• New diagnosis of myeloma (no previous treatment) or a **high suspicion** of a myeloma diagnosis<br>• Eligible for intensive first-line chemotherapy (with or without autologous stem cell transplant) as per up to date national guidelines<br>• Patients local to Sheffield (able to attend for additional research appointments)<br>• Patients planned for follow-up appointments at Sheffield Teaching Hospitals NHS Foundation Trust<br>• Able to give informed consent<br>• Able to tolerate CT scan imaging | • Patients unable or unwilling to give consent<br>• Patients who are too unwell to be recruited<br>• Patients who have had recent chemotherapy for a different malignant condition<br>• Patients with anaphylactic allergy to local anaesthetic<br>• Patients with a known bleeding disorder causing potentially increased risk undergoing additional bone marrow biopsies |

*Note: At recruitment, potential participants will be in the diagnostic work up. If, following consenting, the diagnostic bone marrow biopsy (which will include research samples) confirms an alternative diagnosis to myeloma (e.g. MGUS), the participant will be withdrawn from the study at this point. These patients will not be counted in the final target pilot recruitment numbers and will not proceed to baseline CT imaging.*

**Figure 2.**
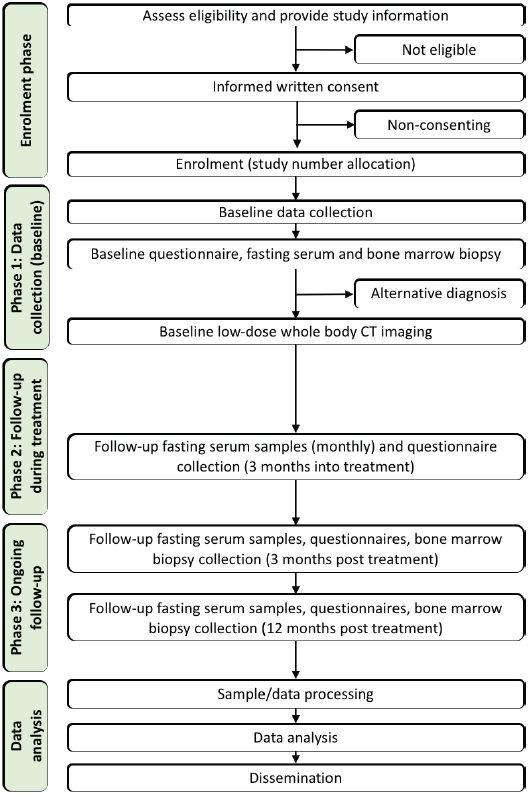
Flow diagram of study design. Following screening and enrolment, patients will receive baseline research investigations prior to initiating first line treatment. Follow-up will include monthly fasting serum analysis throughout, with additional key timepoints of 3 months post treatment initiation, and 3 and 12 months post first line treatment completion. Data will then be processed, analysed and disseminated.

### Study Objectives

#### Co-primary objectives

- *Objective 1*. To evaluate macro-architectural skeletal change using serial cross-sectional WBLDCT imaging (assessment of osteolytic lesions) comparing baseline and follow-up timepoints post first-line treatment for myeloma.
- *Objective 2*. To evaluate micro-architectural changes in micro-CT imaging of bone marrow trephine serial samples (assessment of trabecular bone) comparing baseline and follow-up timepoints post first-line treatment for myeloma.

#### Secondary objectives

1. *Objective 3*. To evaluate histomorphological and immunohistochemical changes in bone marrow trephines, comparing baseline and follow-up post first line treatment of myeloma.
2. *Objective 4*. To evaluate changes in serum bone turnover markers (BTMs: carboxy-terminal collagen crosslinks-1 (CTX-1), pro-collagen type 1 N-terminal pro-peptide (P1NP) and Sclerostin) in myeloma patients at baseline, throughout treatment and at follow-up.
3. *Objective 5*. To evaluate changes in QOL and functional status of myeloma patients at baseline, throughout treatment and at follow-up post first line treatment.
4. *Objective 6*. To assess correlations in bone architecture/microarchitecture and serum BTMs.
5. *Objective 7*. To assess correlations in bone architecture/microarchitecture, with changes in the bone microenvironment (osteoblast and osteoclast number) as evidenced on bone marrow trephine analysis.
6. *Objective 8*. To assess correlations in bone architecture/microarchitecture, with QOL and function status of patients.

### Study Procedures

#### Objectives 1, 6, 7 and 8: Characterization of bone macro-architecture in myeloma patients

Patients who consent to participate will attend for WBLDCT imaging at baseline, and both 3 and 12 months post ASCT (or completion of first line therapy if ASCT did not take place) (Fig. 3). Research images will complement routine imaging taking place at STH (skeletal survey and/or whole body MRI imaging anticipated to become standard of care during the study recruitment period). All research imaging will be reported by an allocated consultant radiologist with specialist interest and experience with diagnostic imaging in myeloma. Reporting will follow a standardised reporting protocol.

**Figure 3.**
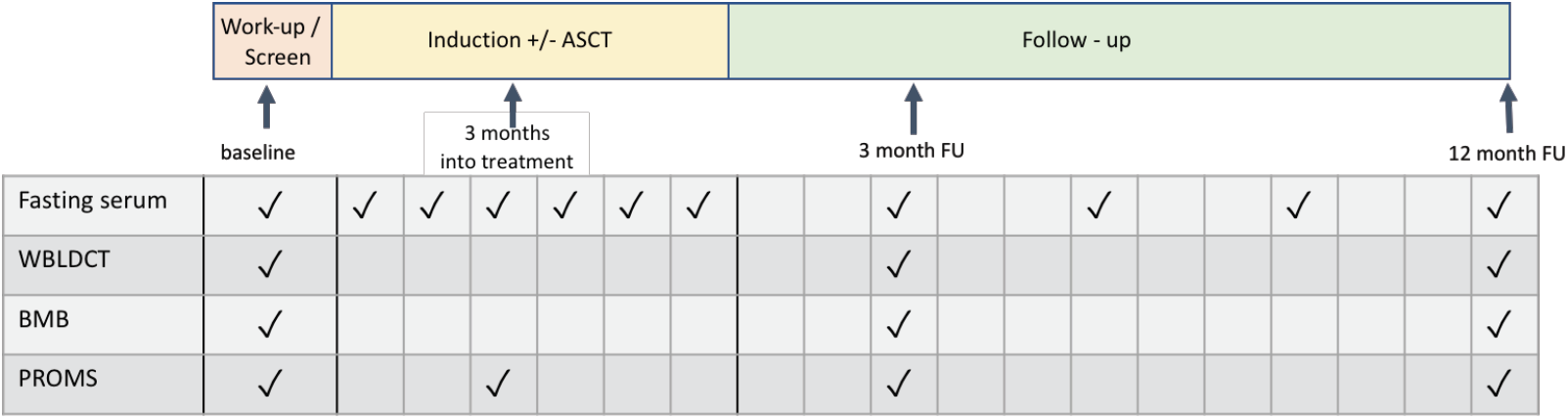
Study follow-up and investigation summary. Research investigations will take place at baseline, throughout induction chemotherapy (+/-ASCT) and for a 12 month follow-up period. *ASCT = autologous stem cell transplant, WBLDCT = whole-body low dose CT, BMB = bone marrow biopsy (aspirate and trephine), fasting serum (for bone turnover markers (BTMs)), PROMS = patient reported outcome markers*.

#### Objectives 2, 3, 6, 7 and 8: Characterization of bone micro-architecture and BMME in myeloma patients

Patients who consent to participate will attend for bone marrow biopsy (aspirate and bone marrow trephine) at baseline, and both 3 and 12 months post ASCT (or completion of first line therapy if ASCT did not take place) (Fig. 3). Research bone marrow trephines will be fixed in formalin and undergo micro-CT imaging using Bruker (Skyscan 1272) scanner with the following protocol: tube voltage 50kV, current 166µA, field of view (FOV) 2016 x 1344, 0.25 Aluminium filter, resolution 10µm, with rotation 0.7(deg) at exposure 832 (ms). Images will be reconstructed and bone volume quantified. Trephines will then be decalcified, embedded within wax paraffin and sectioned for histological analysis. Haematoxylin and eosin (H+E) and tartrate-resistant acid phosphatase (TRAP) staining will be performed, with assessment of osteoblast and osteoclast cell number and distribution. Osteocytes will be assessed by Ploton Silver nitrate staining as previous described. Bone marrow aspirate samples will be processed for CD138 positive cells, then stored at −80°C in RNAprotect for later analysis.

#### Objectives 4 and 6: Quantification of Serum BTMs

Morning fasting serum samples will be collected from study participants at baseline and numerous follow-up timepoints (Fig. 3). Samples will be stored at −80°C and at the end of each recruitment phase, all samples will be run at the same timepoint to avoid unnecessary variability. Bone resorption and bone formation (CTX-1 and P1NP respectively) will be assessed by enzyme-linked immunosorbent assays, as well as serum sclerostin.

#### Objective 5 and 8: Characterisation of changes in QOL throughout first line myeloma treatment

Patients who consent to participate will receive two standardised questionnaires at four timepoints: baseline, completion of first-line treatment, 3 and 12 months post first-line treatment (Fig. 3). The questionnaires used will be EORTC QLQ-C30 and EORTC MY20 (described below).

1. EORTC QLQ-C30 (European Organization for the Research and Treatment of Cancer Quality of Life Questionnaire-Core 30): assessment of QOL in cancer patients. This widely used and validated questionnaire measures global health status, functional scales (physical, emotional, cognitive and social functioning) and symptom scales to assess the effect of cancer on QOL (7).
2. EORTC MY20 (European Organization for the Research and Treatment of Cancer Quality of Life Questionnaire-myeloma module 20): this is a supplementary questionnaire alongside the EORTC QLQ-C30 for assessment of disease symptoms, side effects, future perspective and perceptions of body image. This is widely used and validated in conjunction with EORTC QLQ-C30 to compare QOL between MM patients (8).

Patients will be given the questionnaires during routine hospital visits and asked to return these after completion (with a stamped addressed envelope provided). If a patient feels unable to answer the questionnaires independently, a structured interview (researcher will ask the questions and make note of answers) will be offered.

#### Objectives 6, 7, 8: Correlation Analyses

Bone macro and micro-architectural changes, BTMs and QOL questionnaire results will be correlated and compared between baseline, 3 and 12 month follow-up (as outlined in Fig. 3).

### Sample Size

Following discussion with clinical statisticians, formal power calculations are not deemed possible due to the exploratory nature of this study. A sample size of 20 participants was determined for the pilot study (recruitment cohort 1), based on a realistic predictive timeframe for expected recruitment and study completion within the local area, as well as taking into consideration project costs with the funding available. It is likely that a proportion of participants recruited into the study (recruitment cohorts 1 and 2) will not have a myeloma diagnosis on baseline bone marrow biopsy (confirmation of alternative diagnosis). These patients will not continue to WBLDCT imaging and will exit the study at this time. Based on historical experience, approximately 50 new myeloma diagnoses are anticipated within this centre on a yearly basis, and it is predicted that approximately 20-30 of these would be eligible to participate in this study. Study power calculations will be performed prior to commencing recruitment cohort 2. These calculations will be based on results from recruitment cohort 1 ensuring a more defined (less-exploratory) endpoint for recruitment cohort 2.

## Discussion

MBD associated symptoms (e.g. bone pain) significantly impact the lives of myeloma patients (9), and often remain a chronic feature of the disease, despite good response to cancer-targeting treatments. This, in part, is due to the suboptimal response of bone to recover after lowering tumour burden, with some data supporting the hypothesis that osteoblast function remains altered beyond the removal of inhibitory factors (10). The late-effect burden affecting myeloma patients (11) and the ultimate impact on QOL are becoming a pressured priority. However, bone related outcomes are often overlooked in clinical interventional studies targeting tumour. There remains a clinical need to advance therapeutic options specifically for bone disease, to reduce the burden of this phenomenon on patients and health care resources. Ashcroft *et al*. performed analysis of SREs in myeloma patients and found that 36.7% of reported SREs required a hospital inpatient stay, with the average inpatient period being 20.6 days (12). This data does not include chronic pain associated outcomes, which further increase healthcare resources associated with MBD.

Obstacles to moving this important field forward include the lack of understanding about the state of MBD through current standard of care therapies. There are also challenges in utilising meaningful outcome markers to represent true improvements in bone health for MBD patients. The osteoporosis field tend to focus on bone mineral density and BTM end-points to assess response to treatment, yet in myeloma it is difficult to determine whether this will directly translate to improved patient symptoms and/or clinical outcomes (such as reduced SREs, that require large study populations).

The aim of this study is to provide a comprehensive overview of the BMME as patients undergo first line induction chemotherapy, to try to fully comprehend the ability of bone to “reset” and explore windows of opportunity to successfully interrogate the BMME with novel therapies. Having observed promising outcomes when assessing response to induction chemotherapy in preclinical MBD murine models, we are confident that MBD has the potential to be, at least in part, reversed to improve clinical outcomes (5, 6).

In this study, we will be utilising WBLDCT scans to assess gross skeletal changes, in pathological fracture as well as osteolytic lesion size and number. These findings will be reported descriptively, as well as providing quantified changes in lesion number and size, to assess clinical improvement. A study by Hinge et al. prospectively assessed bone health throughout a phase II interventional study assessing doxorubicin, cyclophosphamide, bortezomib, dexamethasone and lenalidomide (ACVDL) (13) in Denmark (13). In this study, 18 patients receiving first line treatment in the presence of a “target lesion” (osteolytic lesions at least 10mm in size) underwent whole-body low-dose CT imaging (with bone single photon emission CT (SPECT)) and BTM analysis (CTX and P1NP) at baseline, interim (post 4 cycles of chemotherapy), end of treatment and follow-up timepoints. Results demonstrated significant intra-participant variation in osteolytic lesion response, but nonetheless, partial healing was observed in at least one lesion in 7 of the 18 participants (13). This was a higher rate of bone lesion healing than had previously been anticipated, and was thought in part to be due to the use of bortezomib in the treatment regime, which has demonstrated to support bone anabolic activity (14, 15). A retrospective study by Schulze *et al*. assessed retrospective whole-body low-dose multi-detector computed tomography imaging in bortezomib-receiving patients, and demonstrated a 17% rate of radiological sclerosis. They also commented on significant intra-patient variability (16), as has been observed in the VISTA trial post-hoc analysis (17). Our study is also going to be assessing WBLDCT at multiple time points, but our additional analysis will also contribute information about the local BMME.

In the BRAT study, we will aim to assess BTMs (monthly) until end of treatment is reached. We know from the evidence base in osteoporosis diagnosis and monitoring, that BTM levels can change rapidly after initiation of treatment (18). For example, denosumab treatment can significantly reduce CTX within just 24 hours, and zoledronic acid starts to reduce CTX by 2 weeks (19). In the aforementioned study by Hinge *et al*., a significant decrease in P1NP from baseline to interim was observed, followed by an increase towards the end of treatment, with CTX having a rapid reduction after initiation of treatment (13). In assessing BTMs 4 months after chemotherapy was initiated, it is possible that the study team have missed the initial serological changes in bone turnover (18). Other studies have looked at the effect of bortezomib on BTMs, and have demonstrated increased bone formation (14, 15) and reduced bone resorption markers (14, 20, 21).

In our study we will assess iliac crest biopsies (trephine samples) for translatable outcomes of MBD, to include assessment of micro-architecture of the trabecular bone structures, and histological analysis of osteoclast and osteoblasts cells. This should answer questions on how bone turnover alters over the treatment course, including whether bone volume is recovered when quantifying trabecular bone parameters in specific regions of interest. A study by Giuliani *et al*. assessed myeloma patients receiving monotherapy bortezomib for changes in BTMs and trephine biopsy histology, and were able to demonstrate that bortezomib “responders” (as assessed by tumour burden), had significantly increased osteoblast number compared to non-responders (22). We hope to also assess osteoclast and osteoblast number (histologically), as well as bone trephine volume changes (using micro-CT trephine structure analyses).

Overall, this observational study should provide us with an opportunity to assess a number of outcomes markers, which will help us tailor our selection of the most meaningful trial endpoints for future MBD interventional studies moving forward.

### Study limitations

The first phase of this study is a pilot (recruitment cohort 1), and is exploratory in nature, therefore formal power calculations for sample size were not possible. In addition, the single centre approach may limit our recruitment pick up. As this study aims to recruit participants who are eligible for first line induction therapy, frailer (possible older) patients are less likely to be eligible. This is a bias to the study, however, it is one that was deemed necessary as we are keen to assess BMME changes after intensive treatment. It will also allow us to recruit participants who will be undergoing similar myeloma treatment. Another possible bias in this study, is that only English-speaking patients are asked to participate, as the resources are not available to provide translated study documents.

A further limitation to the study is that we cannot accurately predict what other myeloma research studies may be available for patients at the time of recruitment. It is anticipated that there will be a trial assessing first line therapy, and patients will be offered participation in any national trials available. Participation in an interventional trial does not make you ineligible to take part in this observational study. However, it may mean that our recruited cohort will have more varied treatment courses than was initially anticipated in the design of this study. This is unavoidable, but will have to be considered when analysing data.

The requirement of participants to fast for their serum samples may also reduce the acceptability of the study to patients. For some patients, for example poorly controlled diabetes, fasting serum collection will not be feasible. Depending on chemotherapy side-effects, fasting may also have to be reconsidered for some participants.

## Data Availability

No original data is included in this manuscript.

## Ethics and Dissemination

The study is conducted in agreement with the Declaration of Helsinki and local regulations. Potential participants will be given both verbal and written information regarding the study, and appropriate time to consider whether they wish to participate. Written informed consent is obtained from each participant by a member of the research team. Consent forms are filed in a secure location in the site file, and a copy given to the participant for their personal records.

All personal data will be handled confidentially and abide by current data protection regulations. Outside of normal hospital procedures, personal data will only be available to members of the designated research study team. Study participants will be assigned a unique personal study number to pseudo-anonymise the data set. An Excel spreadsheet holding participants identification keys will be stored in a password-protected NHS computer and backed up on a secure, encrypted NHS shared drive. Hard copies of questionnaires, including the site file and consent forms, will be stored in a secure location in a locked office, in a building protected by security.

Results will be disseminated by publication in peer-reviewed scientific journals and participation in scientific conferences. A lay summary of results will be available to study participants.

## Supplementary Materials

Nil

## Author Contributions

**REA;** Conceptualisation, methodology, visualisation, formal analysis, data curation, investigation, writing – original draft preparation, review and editing, project administration, funding acquisition. **IJ;** methodology, formal analysis, data curation, writing – review and editing. **JEB;** supervision, writing – review and editing, funding acquisition. **MAL;** supervision, writing – review and editing, **ADC;** conceptualisation, methodology, investigation, supervision, project administration, resources, funding acquisition. All authors have read and agreed to the published version of the manuscript.

## Institutional Review Board Statement

The study was conducted according to the guidelines of the Declaration of Helsinki, and approved by the Health Research Authority and Health and Care Research Wales (HCRW), and the Yorkshire & The Humber – Bradford Leeds Research Ethics Committee, United Kingdom (REC 18/YH/0275).

## Informed Consent Statement

Informed consent is obtained from all subjects involved in the study.

## Data Availability Statement

No new data were created or analysed in this study. Data sharing is not applicable to this article.

## Acknowledgments

The authors thank Weston Park Cancer Charity (RCN 509803-2) for funding REAs clinical research fellowship, and for providing the support of the initial pilot study (Grant no. R2L). The authors would also like to thank all patients who took the time to consider taking part in this study, as well as staff at the Royal Hallamshire Hospital (Sheffield Teaching Hospitals NHS Foundation Trust) for their support in patient recruitment.

## References

1. Coleman RE. Metastatic bone disease: clinical features, pathophysiology and treatment strategies. Cancer Treat Rev. 2001;27(3):165–76.

2. Snowden JA, Ahmedzai SH, Ashcroft J, D’Sa S, Littlewood T, Low E, et al. Guidelines for supportive care in multiple myeloma 2011. Br J Haematol. 2011;154(1):76–103.

3. Ramsenthaler C, Osborne TR, Gao W, Siegert RJ, Edmonds PM, Schey SA, et al. The impact of disease-related symptoms and palliative care concerns on health-related quality of life in multiple myeloma: a multi-centre study. BMC Cancer. 2016;16:427.

4. Andrews RE, Brown JE, Lawson MA, Chantry AD. Myeloma Bone Disease: The Osteoblast in the Spotlight. J Clin Med. 2021;10(17).

5. Green AC, Lath D, Hudson K, Walkley B, Down JM, Owen R, et al. TGFbeta Inhibition Stimulates Collagen Maturation to Enhance Bone Repair and Fracture Resistance in a Murine Myeloma Model. J Bone Miner Res. 2019;34(12):2311–26.

6. Paton-Hough J, Tazzyman S, Evans H, Lath D, Down JM, Green AC, et al. Preventing and Repairing Myeloma Bone Disease by Combining Conventional Antiresorptive Treatment With a Bone Anabolic Agent in Murine Models. J Bone Miner Res. 2019;34(5):783–96.

7. Aaronson NK, Ahmedzai S, Bergman B, Bullinger M, Cull A, Duez NJ, et al. The European Organization for Research and Treatment of Cancer QLQ-C30: a quality-of-life instrument for use in international clinical trials in oncology. J Natl Cancer Inst. 1993;85(5):365–76.

8. Stead ML, Brown JM, Velikova G, Kaasa S, Wisloff F, Child JA, et al. Development of an EORTC questionnaire module to be used in health-related quality-of-life assessment for patients with multiple myeloma. European Organization for Research and Treatment of Cancer Study Group on Quality of Life. Br J Haematol. 1999;104(3):605–11.

9. Diaz-delCastillo M, Chantry AD, Lawson MA, Heegaard AM. Multiple myeloma-A painful disease of the bone marrow. Semin Cell Dev Biol. 2021;112:49–58.

10. D’Souza S, del Prete D, Jin S, Sun Q, Huston AJ, Kostov FE, et al. Gfi1 expressed in bone marrow stromal cells is a novel osteoblast suppressor in patients with multiple myeloma bone disease. Blood. 2011;118(26):6871–80.

11. Snowden JA, Greenfield DM, Bird JM, Boland E, Bowcock S, Fisher A, et al. Guidelines for screening and management of late and long-term consequences of myeloma and its treatment. Br J Haematol. 2017;176(6):888–907.

12. Ashcroft J, Duran I, Hoefeler H, Lorusso V, Lueftner D, Campioni M, et al. Healthcare resource utilisation associated with skeletal-related events in European patients with multiple myeloma: Results from a prospective, multinational, observational study. Eur J Haematol. 2018;100(5):479–87.

13. Hinge M, Andersen KT, Lund T, Jorgensen HB, Holdgaard PC, Ormstrup TE, et al. Bone healing in multiple myeloma: a prospective evaluation of the impact of first-line anti-myeloma treatment. Haematologica. 2016;101(10):e419–e22.

14. Terpos E, Christoulas D, Kastritis E, Katodritou E, Papatheodorou A, Pouli A, et al. The combination of lenalidomide and dexamethasone reduces bone resorption in responding patients with relapsed/refractory multiple myeloma but has no effect on bone formation: final results on 205 patients of the Greek myeloma study group. Am J Hematol. 2014;89(1):34–40.

15. Heider U, Kaiser M, Muller C, Jakob C, Zavrski I, Schulz CO, et al. Bortezomib increases osteoblast activity in myeloma patients irrespective of response to treatment. Eur J Haematol. 2006;77(3):233–8.

16. Schulze M, Weisel K, Grandjean C, Oehrlein K, Zago M, Spira D, et al. Increasing bone sclerosis during bortezomib therapy in multiple myeloma patients: results of a reduced-dose whole-body MDCT study. AJR Am J Roentgenol. 2014;202(1):170–9.

17. Delforge M, Terpos E, Richardson PG, Shpilberg O, Khuageva NK, Schlag R, et al. Fewer bone disease events, improvement in bone remodeling, and evidence of bone healing with bortezomib plus melphalan-prednisone vs. melphalan-prednisone in the phase III VISTA trial in multiple myeloma. Eur J Haematol. 2011;86(5):372–84.

18. Eastell R, Pigott T, Gossiel F, Naylor KE, Walsh JS, Peel NFA. DIAGNOSIS OF ENDOCRINE DISEASE: Bone turnover markers: are they clinically useful? Eur J Endocrinol. 2018;178(1):R19–R31.

19. Black DM, Reid IR, Cauley JA, Cosman F, Leung PC, Lakatos P, et al. The effect of 6 versus 9 years of zoledronic acid treatment in osteoporosis: a randomized second extension to the HORIZON-Pivotal Fracture Trial (PFT). J Bone Miner Res. 2015;30(5):934–44.

20. Mohty M, Malard F, Mohty B, Savani B, Moreau P, Terpos E. The effects of bortezomib on bone disease in patients with multiple myeloma. Cancer. 2014;120(5):618–23.

21. Terpos E, Kastritis E, Ntanasis-Stathopoulos I, Christoulas D, Papatheodorou A, Eleutherakis-Papaiakovou E, et al. Consolidation therapy with the combination of bortezomib and lenalidomide (VR) without dexamethasone in multiple myeloma patients after transplant: Effects on survival and bone outcomes in the absence of bisphosphonates. Am J Hematol. 2019;94(4):400–7.

22. Giuliani N, Morandi F, Tagliaferri S, Lazzaretti M, Bonomini S, Crugnola M, et al. The proteasome inhibitor bortezomib affects osteoblast differentiation in vitro and in vivo in multiple myeloma patients. Blood. 2007;110(1):334–8.

